# Characterizing spinal reflexes evoked by spinal cord stimulation to restore sensation in people with lower-limb amputation

**DOI:** 10.1101/2023.09.08.23295273

**Authors:** Rohit Bose, Ashley N Dalrymple, Devapratim Sarma, Bailey A Petersen, Beatrice Barra, Ameya C Nanivadekar, Tyler J Madonna, Monica F Liu, Isaiah Levy, Eric R Helm, Vincent J Miele, Marco Capogrosso, Lee E Fisher, Douglas J Weber

## Abstract

**Background:** People with lower-limb amputations lack sensory inputs from their missing limb, which increases their risk of falling. We recently demonstrated that spinal cord stimulation (SCS) can restore sensation in the missing lower limb. Previous studies have shown that SCS can affect motor control by exciting spinal reflex pathways after stroke or spinal cord injury. The effects of SCS on spinal reflex activation have not been studied in people with lower-limb amputation. Furthermore, it is unknown if SCS-evoked spinal reflex activation would perturb walking. Therefore, the goal of this study was to characterize SCS-evoked spinal reflexes in people with lower-limb amputation and quantify effects on gait parameters, including step cycle duration and limb alternation symmetry.

**Methods:** We implanted percutaneous SCS electrodes over the lumbosacral enlargement in 3 people with trans-tibial amputation (2 diabetic neuropathy; 1 traumatic) for 28 or 84 days. SCS was delivered to restore sensation in the missing limb during walking based on signals from a pressure-sensing insole in the shoe under the prosthesis. We used electromyography (EMG) to record posterior root-muscle (PRM) reflexes in the residual limb while participants were seated, standing, or walking. We characterized rate-dependent depression and recruitment properties of the PRM reflexes. We used pressure data from instrumented insoles to measure the step cycle duration and limb alternation symmetry with and without SCS.

**Results:** SCS evoked PRM reflexes in the residual limb muscles in all participants, which was confirmed by the presence of rate-dependent depression at stimulation frequencies ≥2Hz. Overall, there was broad activation of residual limb muscles with SCS that varied with the position of the stimulating electrode relative to the lumbar spinal cord. PRM reflexes were also activated during walking, as confirmed by the presence of rate-dependent depression. However, SCS-evoked PRM reflexes did not disrupt gait, with similar step cycle duration or limb alternation symmetry with and without SCS.

**Conclusions:** Restoring sensation in the missing limb using SCS excites spinal reflexes according to the expected rostral-caudal myotomes but does not disrupt the step cycle duration or limb alternation symmetry in people with trans-tibial amputation.

## INTRODUCTION

By the end of 2050, it is estimated that over 3.6 million people in the United States will be living with a lower-limb amputation as a consequence of vascular disease (54%) or trauma (45%) [1,2]. People with a lower-limb amputation encounter significant challenges in maintaining stability while standing and walking, and as a result, are more likely to fall [3–6]. A primary reason for the higher incidence of falls among people with a lower-limb amputation is the lack of somatosensory feedback from the amputated limb and prosthesis [3,4]. Restoring somatosensation in the missing limb can improve stability during standing and walking [7–9].

Spinal cord stimulation (SCS) is an existing clinical neurotechnology that is used to treat chronic pain by stimulating the dorsal column of the spinal cord with electrodes implanted along the midline in the epidural space [10]. Recently, we demonstrated that by implanting SCS leads laterally in the lumbar epidural space, we can evoke sensations in the missing limb of people with a lower-limb amputation [9]. SCS delivered on the lateral aspect of the spinal cord excites axons from proprioceptors (i.e., primary and secondary muscle spindle and Golgi tendon organ afferents) and mechanoreceptors (i.e., Aβ cutaneous afferents) [11]. By exciting these afferent fibers, SCS engages spinal reflex pathways evoking muscle responses known as posterior root-muscle (PRM) reflexes, which can be recorded using electromyography (EMG) [12–14]. PRM reflexes are a compound reflex response resulting from the multi-segmental activation of proprioceptive and cutaneous afferent fibers that synapse onto spinal motoneurons and interneurons [13,15,16].

By engaging spinal reflex pathways, somatosensory feedback from the feet and limbs plays a crucial role in modulating muscle activity during standing and walking [17–20]. Specifically, somatosensory feedback from the legs facilitates limb loading during stance [21,22], regulates the transitions between the swing and stance phases of walking [23], and facilitates corrective responses to perturbations during walking [18,24,25]. Recent studies have shown that SCS can improve motor control by evoking PRM reflexes in people with stroke [26] and spinal cord injury [27]. The effect of SCS on spinal reflexes in people with amputation is not yet understood. While a long-term goal would be to restore meaningful reflex activity to improve motor control in people with amputation, unwanted muscle activity during gait can cause perturbations or falls. A recent study demonstrated that sensory restoration using peripheral nerve stimulation evoked H-reflexes, and that the elicited sensory percepts did not alter the H-reflexes during gait-related postures [28]. This prior work suggests that people with a limb amputation can perceive sensory feedback provided by electrical stimulation without negatively affecting reflex modulation. Therefore, characterizing spinal reflexes evoked by SCS to restore sensation after limb amputation enables us to understand how SCS affects motor pathways and function, particularly when SCS is used to restore sensation.

Here, we characterize the PRM reflexes evoked by SCS in three individuals with transtibial amputation. We compare their step cycle duration and limb alternation symmetry while walking with or without SCS in one participant. We show that SCS evokes PRM reflexes in the leg muscles in close alignment with the expected myotomal organization of the spinal cord and the rostral-caudal arrangement of the SCS electrodes. We also demonstrate that SCS does not alter step cycle duration and limb alternation symmetry during walking.

## METHODS

All procedures were approved by the Institutional Review Board at the University of Pittsburgh and conformed to the Declaration of Helsinki. Experiments with Participant 3 were performed under an Investigational Device Exemption from the United States Food and Drug Administration. Both studies are registered at ClinicalTrials.gov (NCT03027947 and NCT04547582). Participants provided written informed consent prior to their enrollment in the study. No participants had prior experience with SCS. Data collected and reported in this study were part of another study investigating SCS to restore sensations in the missing limb, improve balance and gait function, and reduce phantom limb pain [9].

### Participants

Three individuals with transtibial amputations participated in this study (Table 1). All participants regularly used a non-motorized lower-limb prosthesis to walk for >40 months before implant. Participants 1 and 3 had an amputation due to complications from diabetic neuropathy and were limited community ambulators (determined by the Amputee Mobility Predictor [29]). Participant 2 had a traumatic amputation and was an active community ambulator. The implant duration was 28 days in Participants 1 and 2 and 84 days in Participant 3.

**Table 1.** Demographic information for research participants.

| Participant ID | Age | Gender | Years since amputation | Nature of amputation | Side of amputation | Ambulation level | Implant duration (days) |
| --- | --- | --- | --- | --- | --- | --- | --- |
| 1 | 56-60 | M | 3.5 | Diabetic | Left | Limited community | 28 |
| 2 | 56-60 | M | 7 | Traumatic | Left | Active | 28 |
| 3 | 60-65 | W | 5 | Diabetic | Left | Limited community | 84 |

We included participants in this study if they were between 22 and 70 years of age, had a unilateral transtibial amputation (but were not excluded for toe amputations on the contralateral limb), were at least 6 months post-amputation at the time of implant, had no serious comorbidities that could increase their risk of participating, were not taking anticoagulant drugs, did not have an MRI-incompatible implanted metal device, and did not have implanted electronic devices such as a pacemaker, defibrillator, or infusion pump. We excluded women who were pregnant or breastfeeding. We also excluded people from the 90-day implant study if their hemoglobin A1c level was above 8.0%, due to increased risk of infection associated with hyperglycemia. Each participant’s neurological and physiological health was evaluated by a physician prior to implantation.

### Implanting leads for epidural spinal cord stimulation

We implanted SCS leads percutaneously under local and/or twilight anesthesia in a minimally invasive, outpatient procedure. SCS leads (Fig 1B) were inserted into the dorsal epidural space over the lumbosacral spinal cord using a 14-guage 4-inch epidural Tuohy needle with the participant lying in the prone position. The leads were steered posterior-laterally using a stylet, guided by live fluoroscopy. We aimed to place the electrodes over the caudal lumbosacral enlargement to target the distal dermatomes of the residual limb and missing foot. In Participant 1, we implanted three 16-contact leads (Infinion, Boston Scientific, Marlborough, MA); two were near the T12-L2 vertebral levels and the third targeted the cauda equina. In Participant 2, we implanted two 16-contact leads (Infinion, Boston Scientific, Marlborough, MA) near the T12-L2 vertebral levels. In Participant 3, we implanted three 8-contact leads (Octrode, Abbott Laboratories, Chicago, IL) near the T12-L2 vertebral levels. We connected each lead to an external stimulator. We stimulated intraoperatively and iteratively adjusted the lead placement based on the participants’ verbal report of the location of sensations evoked in the residual limb. We monitored lead location and migration using weekly X-rays for the first 4 weeks (all participants), then twice monthly after that (Participant 3 only) and compared them to the intraoperative fluoroscopic images. In Participant 1, we anchored the leads using sutures to the superficial layers of skin at the exit incisions, however, all three leads migrated caudally throughout the implant duration. Thereafter, in Participants 2 and 3, we anchored the leads to subcutaneous fascia via a small incision to provide additional stability to the electrode placements. At the end of the study, we removed all percutaneous leads and closed all incision points.

**Figure 1.**
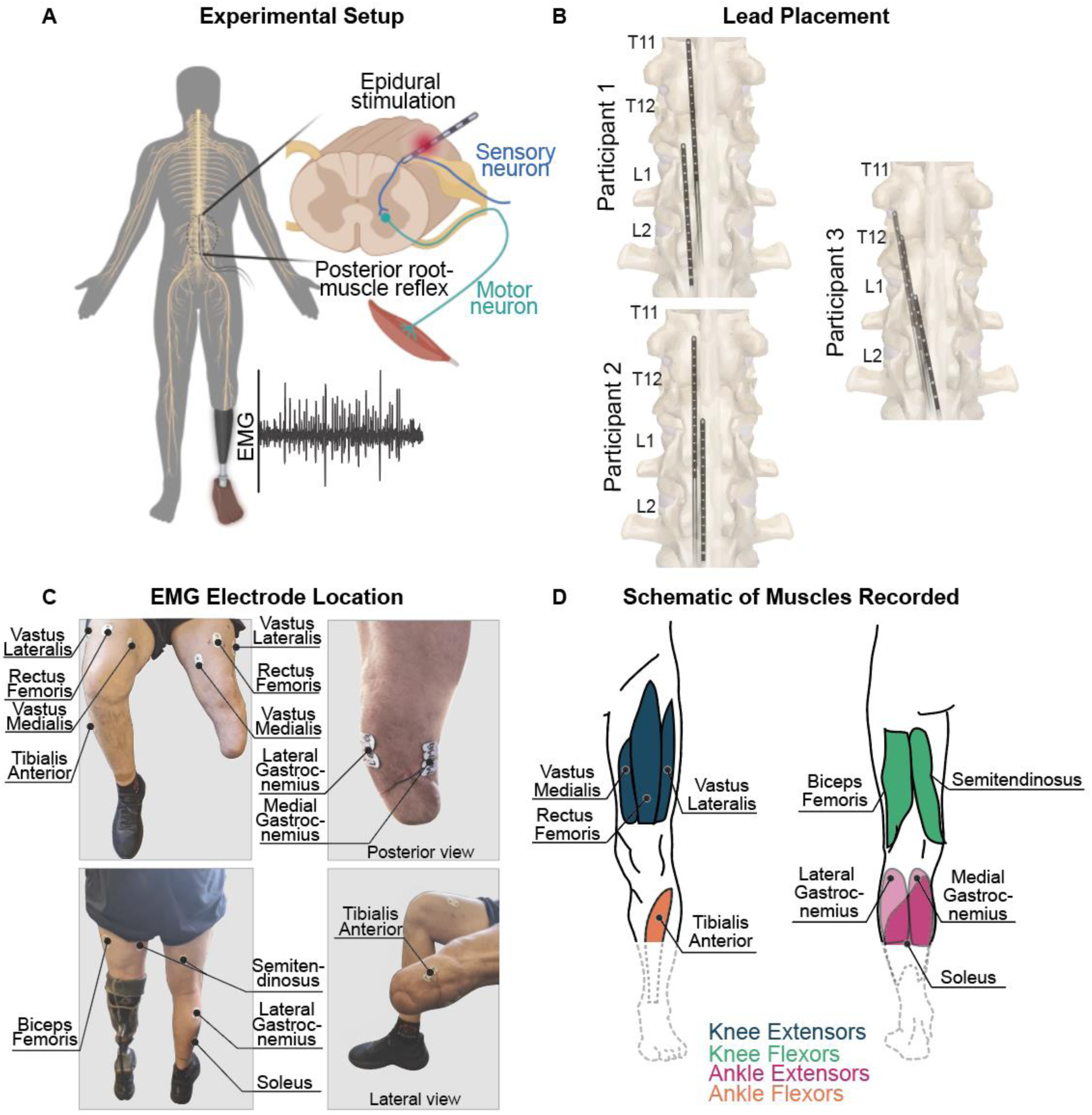
Stimulating and recording setup and electrode locations. (A) Schematic of the experimental setup. Laterally placed epidural spinal cord stimulation (SCS) leads were placed over the lumbosacral region of the spinal cord. Electromyography (EMG) recorded muscle activity and the posterior root-muscle reflex from the muscles of the residual and intact limbs. (B) Approximate location of the SCS electrodes, as confirmed from X-rays, relative to spinal vertebral segments for all participants at week 1 (Participants 2 and 3) and week 2 (Participant 1) post-implant. (C) EMG electrode locations on the intact and residual limbs in Participant 2. (D) Schematic of the muscles where EMG was recorded for all three participants. Functional groupings of muscles are indicated by color: knee extensors (dark blue), knee flexors (dark orange), ankle extensors (light blue), and ankle flexors (light orange).

### Electromyography procedures

EMG data were collected during multiple testing sessions across the implantation period. At the beginning of each day of testing, we cleaned and prepared the skin overlying the muscles of both legs using abrasive gel (Lemon Prep, Mavidon, USA) and alcohol wipes (Nuprep, Weaver, Aurora, CO). We placed bipolar pairs of surface EMG electrodes (Ag|AgCl disposable dual EMG electrodes, MVAP Medical, Thousand Oaks, CA) on the following leg muscles bilaterally in all participants: rectus femoris (RF), vastus Spinal motor mapping by epidural stimulation is (VM), vastus lateralis (VL), biceps femoris (BF), semitendinosus (ST), tibialis anterior (TA), and lateral gastrocnemius (LG) (Figure 1C,D). We placed electrodes on additional ankle extensor muscles of the residual limb: the putative soleus (SO) of Participant 1 and putative medial gastrocnemius (MG) of Participants 2 and 3. For the distal muscles that remained in the socket, we used pairs of soft surface EMG electrodes (Neuroline 700 Series Color Electrodes, MVAP Medical, Thousand Oaks, CA). The putative SO and MG were determined by palpating the residual limb while instructing the participants to attempt plantarflexion while the knee was flexed (SO) or extended (MG). Here, we sometimes refer to the muscles by grouping them according to their function: knee flexors (BF, ST), knee extensors (RF, VM, VL), ankle dorsiflexors (TA), and ankle plantarflexors (LG, MG, SO). We placed a ground electrode (Natus Disposable Ground Electrode, MVAP Medical Supplies, CA) on the anterior superior iliac spine ipsilateral to the amputation.

During testing, Participants 2 and 3 were standing with support (from a walker or assistive frame) or unassisted. Participant 1 had difficulty standing for long durations; therefore, he was either seated or standing, based on his standing tolerance and comfort. We acquired EMG signals at a 30 kHz sampling rate using the Grapevine NIP Data Acquisition System (Ripple, Salt Lake City, UT), with the SurfS2 front-end which has an input range of ±8 mV and 16-bit resolution with 0.25 μV/bit. The SurfS2 includes hardware filters: a 0.1 Hz high pass filter and a 7.5 kHz low pass filter. For participant 3, we used the EMG front-end for the Grapevine system (Sampling rate: 7.5 kHz; Input Range: ± 187.5 mV; ADC: 24-bit resolution with 0.022 μV/bit).

During overground walking, we used wireless data acquisition systems to record EMG signals. In Participant 2, we used the Grapevine NOMAD (Ripple, Salt Lake City, UT) with the SurfS2 front-end. In Participant 3, we used the Trigno Avanti system (Delsys, Natick, MA; sampling rate: 2 kHz; input range: ± 22mV; sensor bandwidth: 10 – 850 Hz).

### Spinal cord stimulation protocol

Electrical stimulation pulses were delivered using three 32-channel constant current stimulators (Nano 2+Stim; Ripple, Salt Lake City, UT). The maximum current output for these stimulators was 1.5 mA per channel. To achieve the higher current amplitudes required for SCS, we built and used a custom circuit board to combine the output of groups of four channels. This increased the maximum current output to 6 mA per channel, with 8 effective channels. We used custom software in MATLAB (MathWorks, Natick, MA, USA) to control stimulation. Stimulation pulses were symmetric and charge-balanced, delivered in a monopolar configuration, with an external return electrode (Natus Disposable Ground Electrode, MVAP Medical Supplies, CA) placed over the right acromion. For Participants 1 and 2, the stimulation pulses were anodic-leading. Due to a change in the software that we used to deliver stimulation, in Participant 3, we delivered stimulation as cathodic-leading. However, due to the biphasic symmetric shape of the stimulus waveform, these changes in polarity did not likely affect recruitment of sensory neurons during SCS [9].

### Study session protocol

Participants attended testing sessions 2–4 days per week throughout the implant period (except week 11 for Participant 3). Within these sessions, reflex and EMG measures were collected between approximately 1 and 3 days per week across all participants. The remaining time was used for quantifying sensory responses to SCS and measuring functional effects of stimulation on balance and gait [9].

#### Detecting SCS-evoked muscle responses

We performed a survey across all SCS electrodes and mapped the evoked muscle responses by delivering 10 pulses at various combinations of stimulation amplitude (2 – 6 mA, in 0.5 – 2 mA steps) and pulse width (0.2 – 0.6 ms, in 0.1 ms steps). For all stimulation parameters tested, we considered an evoked response to be present when the recorded EMG signal exceeded 5 standard deviations (SD) above baseline within 100 ms following a stimulation pulse. We selected pulse widths of 0.4, 0.5, and 0.2 ms for Participants 1, 2, and 3, respectively based on their comfort level.

#### Recruitment curves

We varied the stimulation amplitude through electrodes that evoked sensory percepts from 0.25 mA (below sensory threshold) to the maximum amplitude tolerated by the participants, in increments of 0.25 or 0.5 mA. We repeated stimulation at each amplitude 15 – 20 times at a rate of 1 Hz and randomized the order of each amplitude tested.

#### Confirming reflex responses

If an evoked EMG response is reflex-mediated, (i.e., from activation of sensory afferents), then following two rapid consecutive stimuli, the amplitude of the second response will be smaller in amplitude compared to the first response. This phenomenon is referred to as rate-dependent depression (RDD) [14,30,31]. RDD is not observed when the motoneurons are directly activated by the stimulation pulse. Therefore, we sought to confirm that the recorded responses were indeed reflexive by observing RDD. We stimulated with pairs of pulses with interstimulus intervals of 1s, 500 ms, 200 ms, 100 ms, and 50 ms, corresponding to frequencies of 1, 2, 5, 10, and 20 Hz, respectively. This was done for every amplitude tested for the recruitment curves.

#### Sensory feedback during overground walking

We delivered SCS during overground walking in Participants 2 and 3 starting on Day 23 and Day 22 post-implant, respectively. Due to significant lead migration across weeks in Participant 1, we had to re-map evoked sensations and characterize the PRM reflexes every week. Therefore, we were unable to complete overground walking tests with Participant 1. We instructed the participants to walk at their self-selected speed across a 6-meter walkway. Wireless pressure sensing insoles (Moticon Insole 3, Munich, Germany) measured limb loading. When the plantar pressure of the insole in the prosthetic foot exceeded a pre-programmed threshold to indicate the stance phase, the stimulation was triggered to evoke sensations that emanated from the participant’s missing foot [9]. Therefore, SCS was delivered only during the stance phase of the prosthetic limb. Stimuli in Participant 2 were on-off triggered at a constant amplitude when insole pressure exceeded 35 N/cm^2^. For Participant 3, stimulus amplitude was linearly modulated with insole pressure above a threshold of 3 N/cm^2^. Stimuli were delivered at 50 Hz for Participant 2 and 90 Hz for Participant 3, with a pulse width of 0.2 ms for both participants. These stimulation parameters were chosen because they evoked reliable sensory percepts and were comfortable for the participants.

### Data analysis methods

To allow for comparisons across participants, we express the stimulus intensity as a unit of charge (C), which was calculated by multiplying the amplitude of the cathodic phase of the stimulus (A) by the pulse width (s).

#### Measuring muscle recruitment and reflex responses

EMG data were bandpass filtered from 30 to 800 Hz (4^th^ order Butterworth filter) with a notch filter at 60 Hz. To construct the recruitment curves, for each stimulation charge, we calculated the average peak-to-peak amplitude of the evoked EMG signal.

We measured RDD by comparing the peak-to-peak amplitude of the evoked EMG signal from successive stimuli. We expressed the peak-to-peak amplitude of the evoked EMG signal from later stimulus as a percentage of the peak-to-peak amplitude of the first response. We repeated this procedure across all frequencies (1, 2, 5, 10, and 20 Hz).

We characterized the relative activation of the lower-limb muscles across spinal segments from a 1 µC stimulus pulse for Participant 1, 3 µC for Participant 2, and 1.2 µC for Participant 3. The stimulation charge was selected based on the maximum tolerable charge that could be delivered across a majority of the electrodes. We performed this functional mapping procedure within two days following an X-ray to ensure accurate positioning of the electrodes. The peak-to-peak amplitude of the EMG response from each SCS electrode was standardized using the z-score. We then found the difference between the z-scores for each muscle’s maximal and minimal activation (*Amplitude_diff_*), across each electrode using the following equation:

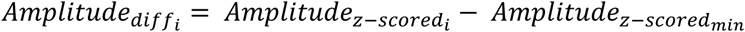

Where *i* denotes individual muscle, 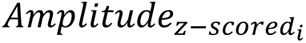 denotes peak-to-peak amplitude of the response (z-scored) of each muscle and 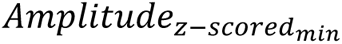 indicates minimum peak-to-peak amplitude of the response (z-scored) across all the muscles and electrodes in a participant. Therefore, an amplitude value of zero indicates the minimum magnitude of muscle activity for that participant across all electrodes.

#### Quantifying muscle activity, step cycle duration, and limb alternation symmetry during walking

Participants 2 and 3 were instructed to walk overground at a self-selected speed with and without SCS applied during the stance phase. We determined the duration of each step cycle as the time from loading onset between steps of the residual limb using heel contact force recorded from the insoles. The participants walked with and without SCS in a single session after receiving SCS for several weeks. For all comparisons, we tested for normality using the Shapiro–Wilk test and assessed the homogeneity of variance using Levene’s test. Step cycle durations with and without SCS were compared using a Wilcoxon signed-rank test in Participant 2, and a t-test in Participant 3. A p-value < 0.05 was considered significant. We performed equivalence testing using the two one-sided t-test (TOST) method using the TOST function in MATLAB [32,33] to compare the similarity in the step cycle durations across conditions in each participant.

We also calculated step alternation symmetry between the residual and intact legs by finding the difference in the time spent loading using the sum of pressure recorded from the insoles [34]. Specifically, we measured the time each leg spent loaded within each step cycle and found the difference between the half-way loading time for each limb. One step cycle was set equal to 360°, and the difference in half-way loading times of each leg was converted to degrees within the step cycle to indicate the alternation. Perfect alternation between the two legs is equal to 180°, indicating that they are exactly out of phase from each other. A limb alternation greater than 180° indicates that the residual limb was loaded for a longer duration (had a longer duration of the stance phase), and a limb alternation less than 180° indicates that the intact limb was loaded for a longer duration. Limb alternation symmetry with and without SCS was compared using a t-test in both participants. The TOST method was used to compare alternation symmetry across conditions in each participant.

EMG from the overground walking experiments were high-pass filtered using a 2^nd^ order Butterworth filter with a cut-off frequency of 300 Hz. We determined the PRM reflex latency and amplitude of consecutive responses across a variety of interstimulus intervals (14 -49 Hz) to measure RDD. The latency was defined as the time from the onset of the stimulation to the onset of the response. The onset of the response was detected when the amplitude exceeded two standard deviations beyond the mean baseline period and was confirmed with manual inspection.

## RESULTS

We observed that SCS evoked responses in both the residual and intact limbs for Participant 1 and 2, and only in the residual limb for Participant 3. The intact limb responses appeared primarily for the contacts located more medial. The thresholds for the residual limb were 0.86 ± 0.2 µC, 1.2 ± 0.55 µC and 1.23 ± 0.31 µC for Participant 1, 2 and 3 respectively. The thresholds for the intact limb were 0.8 ± 0.22 µC in Participant 1 and 1.2 ± 0.56 µC in Participant 2 across all the muscles. However, our goal in this study was to characterize the PRM reflexes evoked in the residual limb; therefore, our analyses are limited to the responses from that limb.

### SCS evokes posterior root-muscle reflexes in the residual limb

In all the three participants, SCS evoked EMG responses in muscles throughout the residual limb. To establish that the responses were reflexive in nature, we quantified RDD across several interstimulus intervals ranging from 50 ms to 1 s, corresponding to frequencies of 20 Hz to 1 Hz, respectively. RDD of the evoked responses was present across muscles in all participants, indicating they were indeed PRM reflexes (Figure 2A,B). At 1 Hz, there was a mixture of small magnitude depression and facilitation (< 20% in most instances; Figure 2B). In all participants, as the interstimulus interval decreased (higher frequency), the peak-to-peak amplitude of the second response decreased compared to the first response. At 5 Hz (Participant 1), ≥ 10 Hz (Participant 2), and ≥ 2 Hz (Participant 3), RDD was present in all muscles in which an evoked response was recorded. At the highest frequency tested (5 Hz for Participant 1, 20 Hz for Participants 2 and 3), the mean (± SD) reduction in the second response compared to the first response was 38.5% (± 8.1%) for Participant 1, 83.6% (± 4.8%) for Participant 2, and 80.14% (± 4.97%) for Participant 3 (Figure 2B). This confirmed that SCS-evoked muscle activity is mediated by the spinal reflex pathway and not from direct activation of motoneurons.

**Figure 2.**
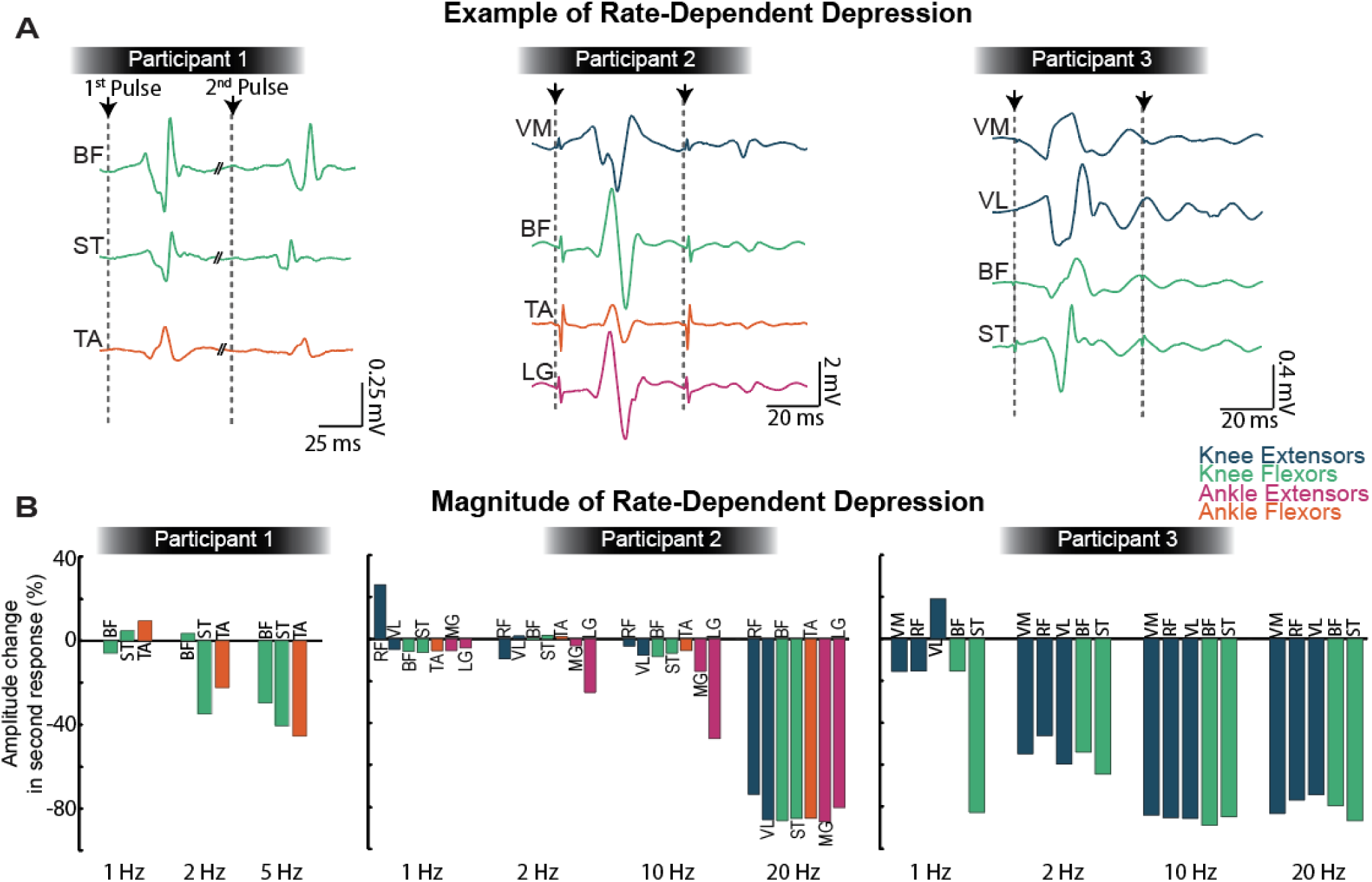
SCS evoked posterior root-muscle (PRM) reflexes. (A) Characteristic PRM reflex responses evoked at 1.5 times reflex threshold with an inter-stimulus interval of 100ms (for Participant 1) and 50ms (for Participant 2 and 3), demonstrating rate-dependent depression (RDD) of the second response compared to the first. (B) The change in the peak-to-peak amplitude of the second response compared to the first response for each muscle where reliable PRM reflexes were evoked in each participant. Muscles are grouped by color: knee extensors (blue); knee flexors (green); ankle extensors (pink); ankle flexor (orange). VL = vastus lateralis, RF = rectus femoris, VM = vastus medialis, BF = biceps femoris, ST = semitendinosus, TA = tibialis anterior, MG = medial gastrocnemius, LG = lateral gastrocnemius.

### Characterizing PRM reflex recruitment

We constructed recruitment curves by varying the stimulating amplitude through electrodes that evoked sensory percepts in each participant and measured the peak-to-peak amplitude of the PRM reflex responses (Figure 3). Nearly all recruitment curves did not reach their saturation point, indicating that the stimulation intensity required to achieve maximal reflex recruitment was greater than 6 mA. Due to limitations in the design of our stimulator, we did not stimulate above 6 mA.

**Figure 3.**
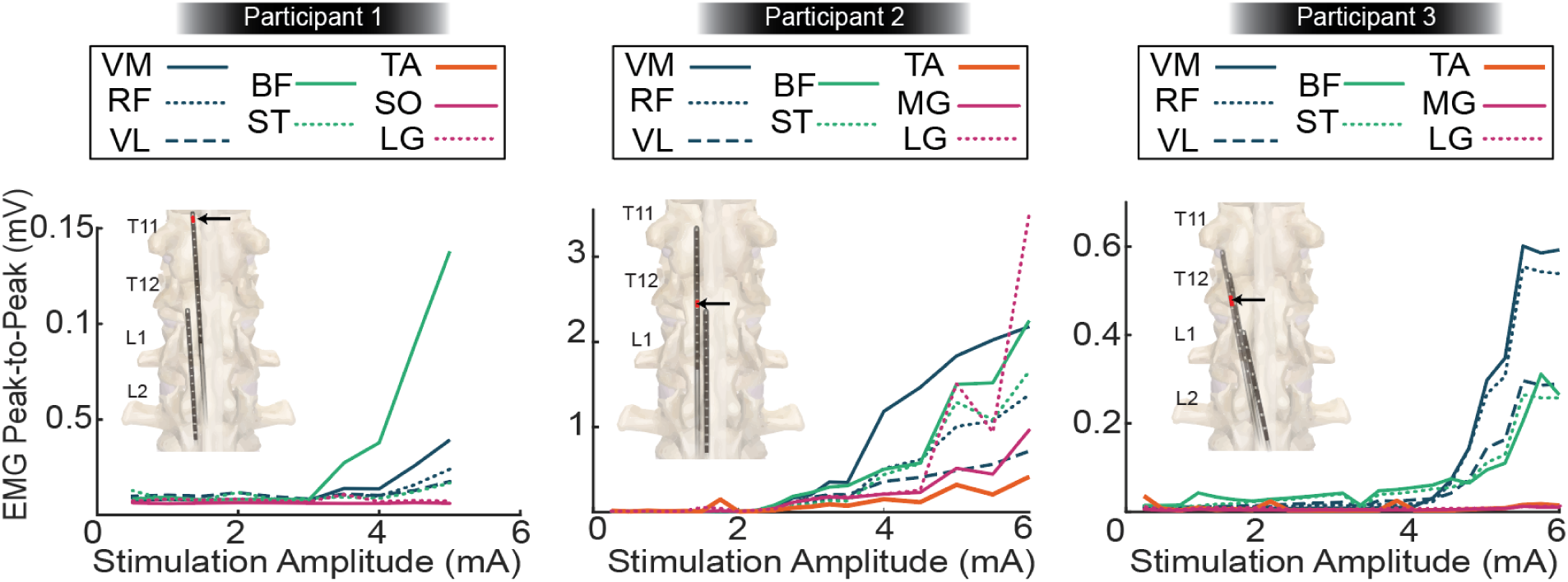
Examples of recruitment curves when stimulation amplitude was increased through an electrode that evoked sensory percepts in the missing limb (electrode indicated by the arrow in inset). Muscles are grouped by color: knee extensors (blue); knee flexors (green); ankle extensors (pink); ankle flexor (orange). VL = vastus lateralis, RF = rectus femoris, VM = vastus medialis, BF = biceps femoris, ST = semitendinosus, TA = tibialis anterior, MG = medial gastrocnemius, LG = lateral gastrocnemius.

Further, we characterized the activation profiles of the residual limb muscles with respect to the rostral-caudal placement (T11 to L1 vertebrae) of the electrodes over the spinal cord. At the most rostral electrode (near the T11 vertebra), there was minimal activation of the most proximal muscles in all participants (Figure 4). Stimulation through more caudal electrodes exhibited diffuse activation of all muscles in all participants. However, the evoked reflex activity was larger for muscles in the myotomes corresponding to the location of the SCS electrode [44]. We did not observe selective activation of an individual muscle in any participant with any electrode, likely due to the partially overlapping distribution of myotomes across different spinal segments.

**Figure 4.**
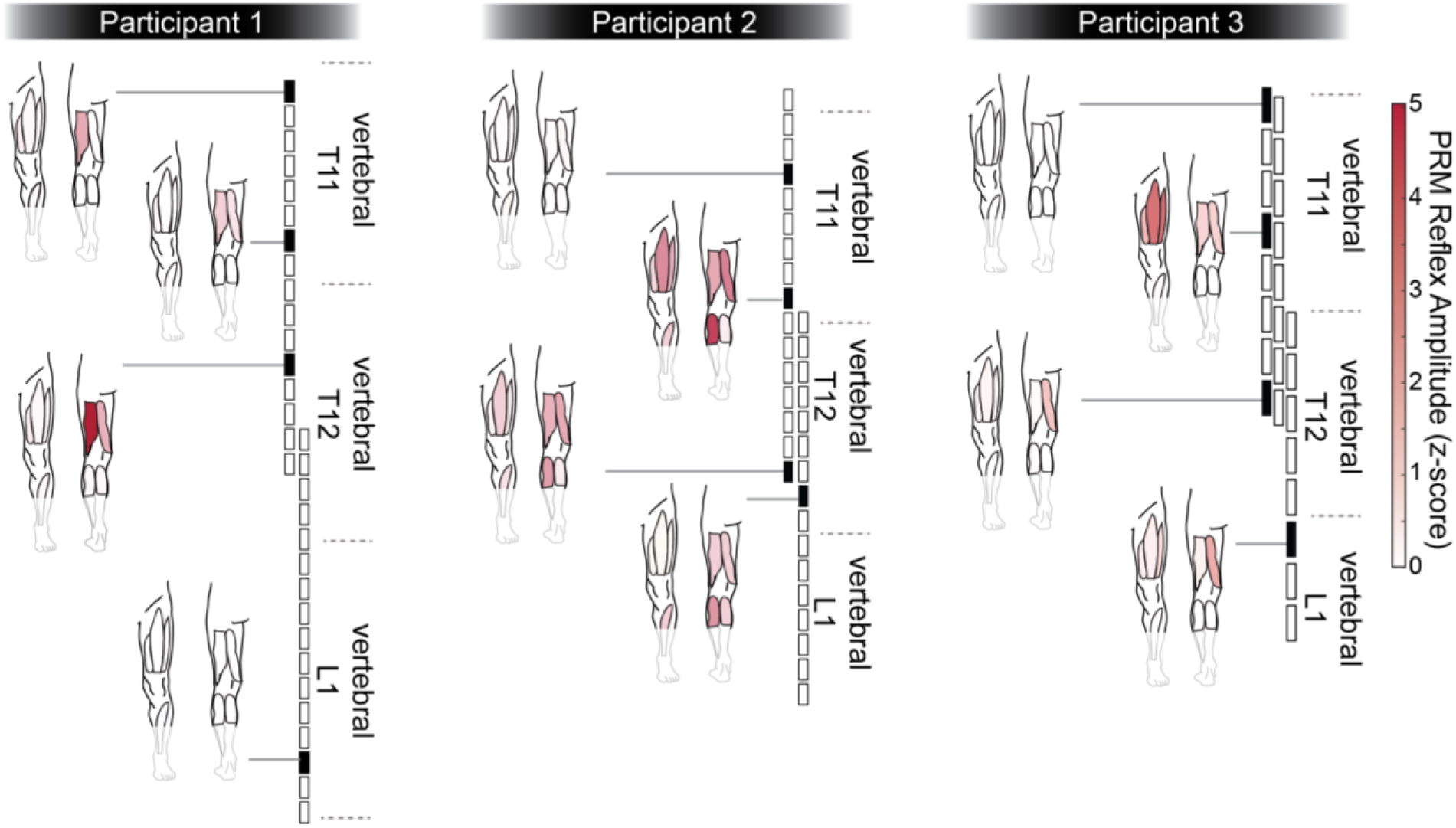
Posterior root-muscle (PRM) reflex peak-to-peak amplitude (z-scored) following stimulation through electrodes according to their rostral-caudal placement over the lumbosacral enlargement.

### PRM reflexes are present during walking

We investigated whether SCS-evoked PRM reflexes were present during overground walking. While Participant 2 was walking with SCS, PRM reflex responses were observable in the residual limb throughout the stimulation train when the interstimulus interval was greater than 20 ms (< 50 Hz). PRM reflexes were detected only in the EMG of VL and RF muscles of the residual limb; the walking EMG data from the other muscles were too noisy to analyze. The onset latency of the reflex response in both VL and RF muscles was 17 ms (Figure 5). If PRM reflex responses were observable following the first two consecutive stimulation pulses at the start of the train, they were analyzed for RDD. This occurred 17 times in the EMG of the VL muscle and 11 times in the EMG of the RF muscle. In all instances of two consecutive reflex responses at the start of a stimulation train, the second response was smaller in amplitude than the first response, confirming the presence of RDD. The amplitude of the second response was 65.2% (± 18.7%) and 66.4% (± 25.8%) smaller than the amplitude of the first response for VL and RF, respectively.

**Figure 5.**
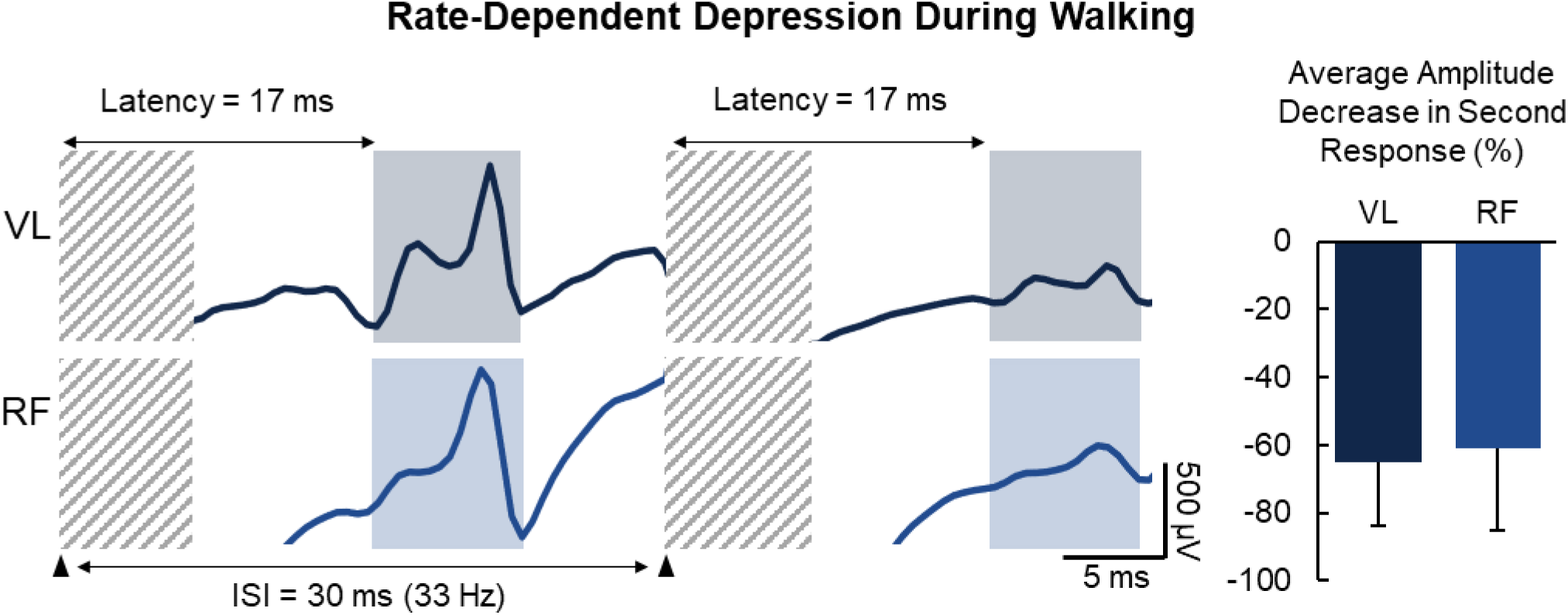
Rate-dependent depression of posterior root-muscle reflexes. (A) Examples in vastus lateralis (VL) and rectus femoris (RF) muscles while Participant 2 walked. Stimulation artefact is covered by gray diagonal lines; PRM reflex is outlined by shaded boxes. (B) Average (‒standard deviation) decrease of the second reflex amplitude compared to the first response. ISI = interstimulus interval.

### SCS does not alter step duration or limb alternation symmetry

Participant 2 walked with a step cycle duration of 1.40 ± 0.09 s without SCS and 1.38 ± 0.10 s with SCS, which were not significantly different (p = 1.0; n = 150 steps; Figure 6A). In fact, the step cycle durations with and without SCS were equivalent within ± 5%. Participant 3 walked with a step cycle duration of 1.08 ± 0.14 s without stimulation and 1.05 ± 0.2 s with stimulation. These cycle durations were also not significantly different (p = 0.66; n = 42 steps total; Figure 6A) and were equivalent within ± 10%.

**Figure 6.**
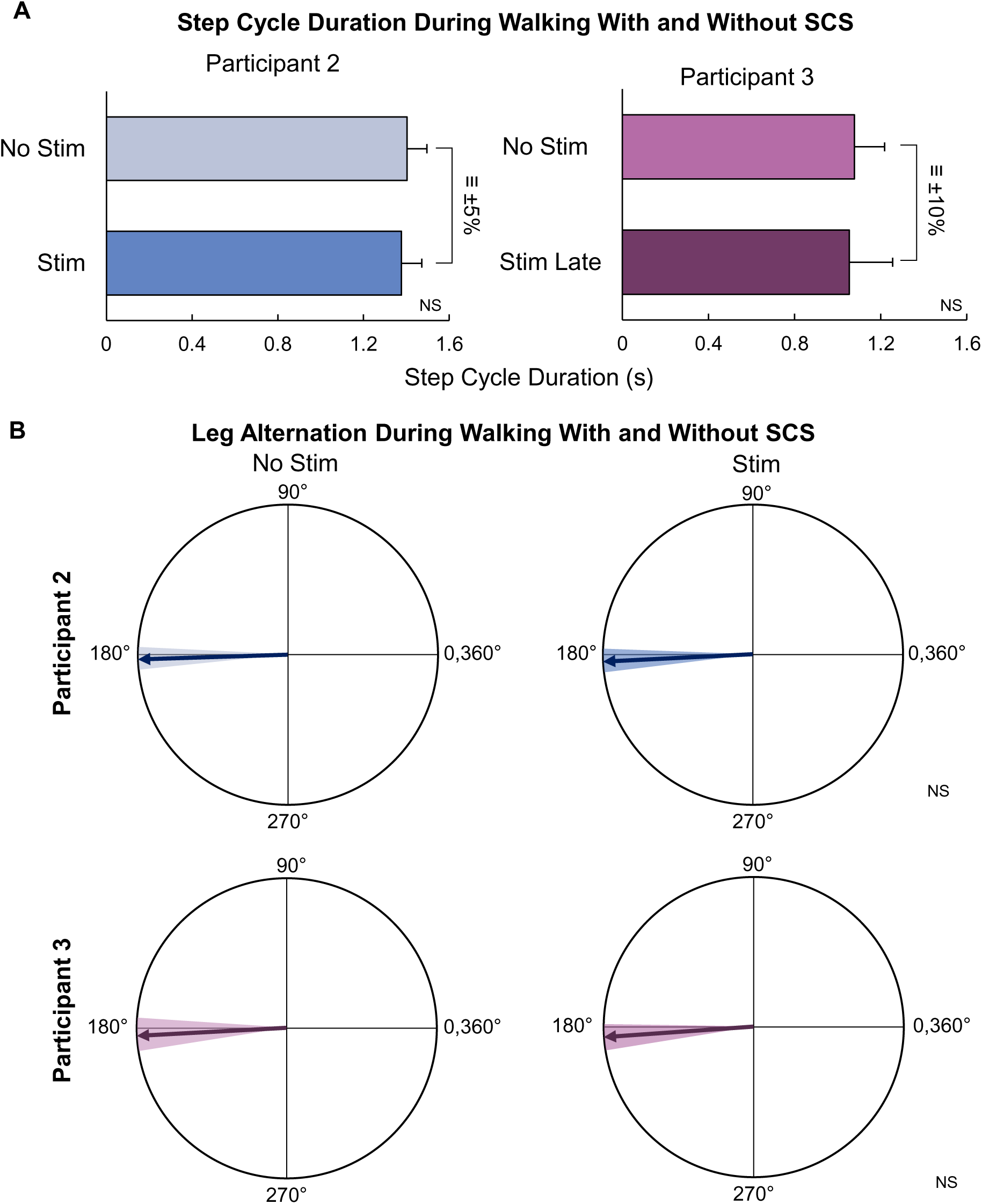
Walking metrics with and without SCS for Participants 2 and 3. (A) Average (+standard deviation) step cycle duration. (B) Limb alternation symmetry. The arrow and shaded area indicate average and standard deviation, respectively. A 180° angle indicates perfect alternation between the two limbs. NS: Not significant; =: equivalent.

Furthermore, SCS did not induce a change in limb alternation symmetry in either participant (Figure 6B). Both Participant 2 and 3 had near perfect limb alternation symmetry without SCS (Participant 2: 181.5° ± 4.4°; Participant 3: 182.9° ± 6.2°). With SCS, the limb alternation symmetries were 182.2° ± 4.5° for Participant 2, and 183.6° ± 5.1° for Participant 3. The limb alternation symmetries with SCS were not significantly different to those without SCS (Participant 2: p = 0.13; Participant 3: p = 0.62), nor were they equivalent in either participant, according to the TOST method. A limb alternation of 180° indicates perfect alternation between the intact and residual limb; therefore, both participants exhibited normal limb alternation without SCS, and this was unaltered with SCS. Similarly, both the residual and intact VL muscles exhibited alternating bursting activity with similar timing during walking without and with SCS (Figure 7).

**Figure 7.**
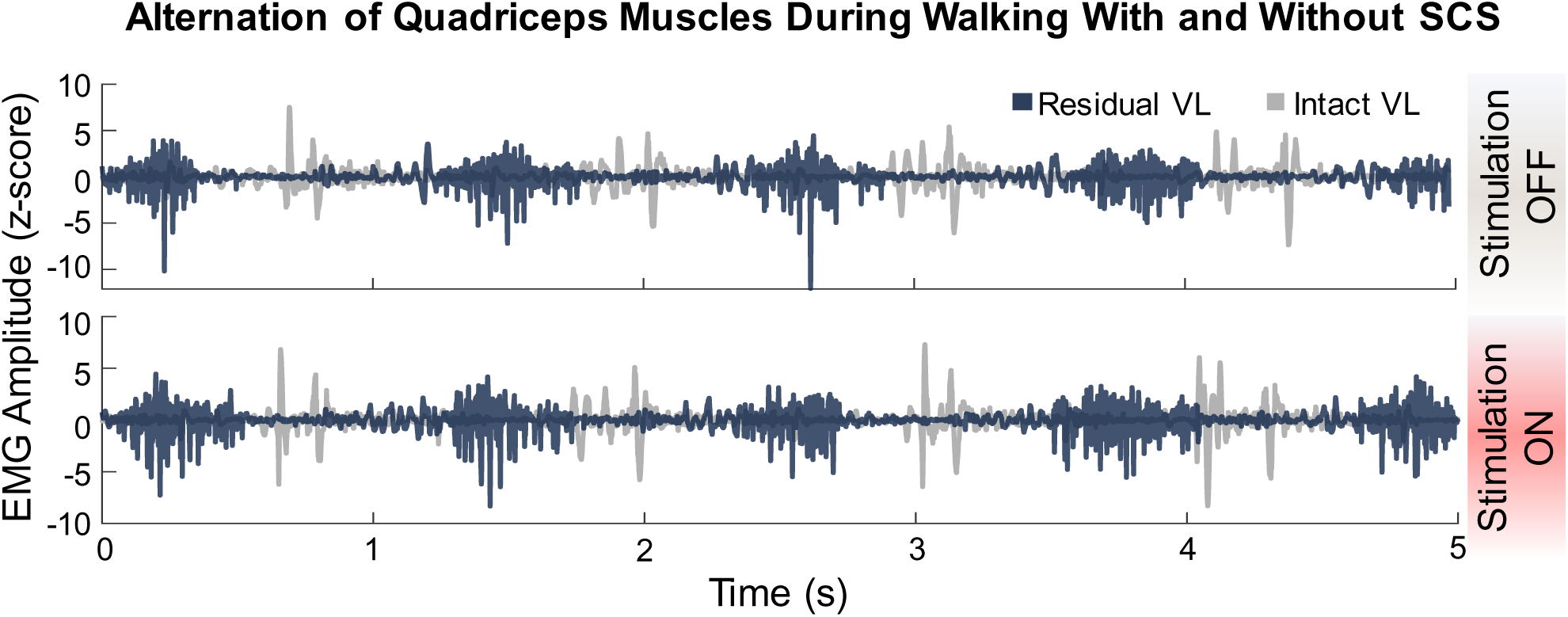
Vastus lateralis EMG during walking. Unfiltered bursts of muscle activity from the alternating residual and intact vastus lateralis (VL) muscles during walking with and without spinal cord stimulation (SCS) in Participant 3 on Day 63.

## DISCUSSION

Our previous work showed that SCS can evoke sensory percepts in a missing limb [9,35,36]. However, sensorimotor networks are highly integrated, especially in the spinal cord, via reflexive pathways. The primary goal of this study was two-fold: (1) to characterize the muscle activity evoked via spinal reflex pathways from SCS, and (2) to investigate whether SCS-evoked reflexes perturbed gait in people with trans-tibial amputation.

### PRM reflexes are present following limb amputation

SCS targets the dorsal spinal roots, exciting large diameter afferent fibers [11,37]. PRM reflexes consist of a superposition of H-reflexes and cutaneous afferent inputs [15,16,38], whereas H-reflexes arise from the excitation of Ia afferent fibers and their synapses onto motoneurons only [39]. Here, we present and characterize PRM reflexes evoked using SCS in people with a lower-limb amputation, with and without diabetic neuropathy. PRM reflexes were confirmed by measuring RDD, which was present in all participants demonstrating that stimulation was not directly activating motor neurons in the ventral spinal cord or roots.

We were able to evoke PRM reflexes in all participants, including two that had diabetic neuropathy. In diabetic neuropathy, tendon tap and Hoffman (H-) reflexes are prolonged or absent [40–42]. The fact that PRM reflexes were evoked consistently in this population demonstrates their utility to probe the spinal cord after amputation, which has implications for future work further studying gait differences and phantom limb pain after lower-limb amputation. The current work is the first step towards the investigation of spinal reflexes after limb amputation.

### Muscle recruitment following limb amputation and SCS

When stimulating at any location in the lumbosacral enlargement, we observed broad recruitment of muscles in the residual limb, with the strongest activation corresponding to the expected myotome for that region of the spinal cord [43–45]. For instance, the strongest activation of the knee extensors was observed at vertebral level T11 whereas the knee flexors were observed at vertebral level T12. The broad recruitment may have occurred because of a number of factors, including: 1) current spread in the cerebrospinal fluid (CSF) causing excitation of multiple spinal root segments [46], 2) the activation of intraspinal networks [34,47], or 3) due to plastic changes in the spinal circuitry following limb amputation [48]. Current spread through the CSF is known to occur with SCS, leading to the activation of multiple spinal root segments [46]. However, a recent study showed similar broad recruitment of muscles from direct stimulation of the lumbosacral spinal roots [43]. Similarly, the activation of intraspinal networks connecting several motoneuron pools throughout the lumbosacral enlargement can contribute to multi-joint movements [47,49–51]. However, the activation of intraspinal networks typically consists of functional synergistic movements, such as flexion or extension synergies. Therefore, it is not clear why there would be broad activation of all lower-limb muscles after limb amputation. A study in non-human primates showed that, following limb amputation, the motoneurons innervating the distal muscles survive, and that they reinnervate different muscle targets in the residual limb [48]. This expansion in the motor unit innervation is a possible reason for the broad muscle recruitment with SCS shown here, especially in Participant 2, who had a traumatic amputation.

### SCS during walking

SCS activates multiple types of afferent fibers simultaneously, including proprioceptive and cutaneous afferents, which excite spinal reflexes [15,16,38,57]. Spinal reflexes can play a role in the modification of timing of gait phases during walking [19]. The role of spinal reflexes on the modification of gait patterns is dependent on which phase of the gait cycle the limb is currently in. For example, activation of the stretch reflex can provide assistance for weight-bearing during stance [21,58]. The Golgi tendon reflex reverses its role from inhibition to excitation during the stance phase in order to contribute to weight-bearing [22,59–61] and can also modify the timing of the step cycle by elongating the stance phase [23,62]. Furthermore, cutaneous reflexes originating from distal peripheral nerve stimulation can modify the timing of the swing and stance phases, depending on when the stimulation is delivered during the gait cycle and the region of the foot innervated by the stimulated nerve [63,64]. Computational modelling studies have also shown that afferent stimulation can modulate the timing of activation of the spinal central pattern generator [65]. The relationship between PRM and other reflexes is not known for certain, but PRM reflexes have been proposed to consist of a superposition of H-reflexes and cutaneous reflexes [15,16,38].

We had posited two possible outcomes that could result from SCS-evoked reflex activity: (i) evoking widespread reflex activity may impede or perturb walking, or (ii) SCS could facilitate useful reflex activity to improve walking. We observed PRM reflexes in the residual limb during walking, indicating that indeed, these sensorimotor reflex pathways were activated by SCS. However, the excitation of afferent fibers and PRM reflexes did not alter the timing of activation of muscles to affect step cycle duration and limb alternation symmetry. Step cycle duration in young, non-injured individuals typically ranges from 0.94 – 1.07 s [52,53]; however, in people with limb amputation, step cycle durations are typically shorter, but vary according to disability and can reach durations similar to non-injured individuals [54–56]. Our participants walked with step cycle durations that were longer or similar to non-injured individuals. Importantly, the step cycle duration was similar with and without SCS. Therefore, we can conclude that SCS with sensory feedback and reflex activation did not hinder their walking speed. Furthermore, the limb alternation symmetry was near 180°, indicating perfect alternation, without SCS, and was unaltered by SCS. Our prior study demonstrated improved balance control and gait stability with sensory feedback from SCS, notably with greater improvements in the most challenging balance and gait conditions [9]. In this work, we specifically investigated features of walking under the simplest condition – during flat overground walking at a self-selected speed. Features such as step cycle duration and limb symmetry were already near-normal, and we sought to determine if sensory feedback from SCS would interfere with the normal aspects of gait. Therefore, we show that our concern that SCS-evoked reflex activity could interfere with walking function was not realized in our participants, similar to the findings from the recent study using peripheral nerve stimulation to restore sensation [28]. Between our prior work and the current study, we suggest that sensory feedback from SCS can improve balance and gait stability during challenging tasks, but SCS-evoked PRM reflexes do not impede already normal gait parameters.

It is possible that input from supraspinal regions dominated over any effects from the spinal reflexes on the gait cycle. Indeed, previous studies have shown that supraspinal inputs modulate proprioceptive afferent input during walking, resulting in different H-reflex and PRM reflex amplitudes throughout the gait cycle [21,66–68]. Volitional control originates in the primary motor cortex, but this region receives projections from the premotor area and supplementary motor area [69]. Additionally, the mesencephalic locomotor region in the brainstem can initiate locomotion as well as modify cadence [70,71]. All these regions and others directly or indirectly project to the spinal cord and are capable of controlling the timing of the gait cycle and modulating spinal reflexes. The interplay between the SCS-evoked PRM reflexes with the supraspinal input and afferent input from the residual limb is still obscure. Further investigation is required to understand the effect of SCS on individual lower-limb muscles during different functional tasks.

### Limitations

The results presented here are from a small sample of individuals; therefore, we present the results for each individual and do not perform group analyses. Future work will aim to further characterize PRM reflexes over time in people with lower-limb amputation receiving SCS for sensory restoration. This will enable us to understand how PRM reflexes are modulated over time and their role in the improvements in challenging balance and gait tasks.

Participant 1 had difficulty standing for extended durations; therefore, much of the sensory testing and PRM reflex data were collected during sitting in a chair. The data for Participants 2 and 3 were collected during standing with or without the assistance from a standing frame or walker. Irrespective of these postural differences, we have shown that SCS could evoke PRM reflexes in all participants. Similarly, other studies using tSCS have shown recruitment of PRM reflexes with varying postures [72,73]. In the future, we aim to systematically characterize the effect of posture on PRM reflexes evoked by SCS.

### Clinical relevance

SCS is a common method to reduce pain, with as many as 50,000 people receiving SCS implants each year to treat chronic pain [74]. Our previous work demonstrated that SCS can be used to restore sensation in the missing limb, improve function during walking, and reduce phantom limb pain [9]. In this study, we show that the SCS used to achieve those effects also engages spinal reflexes, and that the spinal reflexes do not disrupt the timing or symmetry of gait. Therefore, with improved sensorimotor function, over time, SCS may be able to reduce the incidence of falls in people with lower limb amputation. Future studies will focus on additional gait tasks and the role of reflex recruitment and muscle activation on improvements in gait over longer durations of time.

## CONCLUSIONS

SCS to restore sensation in the missing limb of people with trans-tibial amputations evoked PRM reflexes in the residual limb. SCS broadly activated the muscles of the residual limb in all participants. When SCS was delivered to provide sensory feedback during walking, it did not cause unwanted muscle activation nor alter the step cycle duration or limb alternation symmetry.

## Data Availability

All data produced in the present study are available upon reasonable request to the authors

## LIST OF ABBREVIATIONS

SCS: spinal cord stimulation
PRM: posterior root-muscle
EMG: electromyography
RF: rectus femoris
VM: vastus medialis
VL: vastus lateralis
BF: biceps femoris
ST: semitendinosus
TA: tibialis anterior
LG: lateral gastrocnemius
MG: medial gastrocnemius
SO: soleus
SD: standard deviation
RDD: rate-dependent depression
tSCS: transcutaneous spinal cord stimulation
CSF: cerebrospinal fluid

## ACKNOWLEDGMENTS

The authors would like to express our sincerest gratitude to our research participants for their time and dedication to furthering the field of neuroprosthetics. We would like to thank Debbie Harrington, Casey Konopisos, and Alayna Schwerer for their assistance with the IRB protocol, clinical trial registration, IDE application, and participant recruitment. Figure 1A was created using BioRender.com

## CLINICAL TRIAL INFORMATION

This study was registered under clinical trials NCT03027947 and NCT04547582.

